# Establishment and validation of pre-therapy cervical vertebrae muscle quantification as a prognostic marker of sarcopenia in head and neck patients receiving definitive cancer surgery

**DOI:** 10.1101/2021.08.26.21262353

**Authors:** Brennan Olson, Jared Edwards, Catherine Degnin, Nicole Santucci, Michelle Buncke, Jeffrey Hu, Yiyi Chen, Clifton D. Fuller, Mathew Geltzeiler, Aaron J. Grossberg, Daniel Clayburgh

## Abstract

**Importance:** Sarcopenia, or diminished skeletal muscle mass, is prognostic for survival in patients with head and neck cancer (HNC). However, identification of this high-risk feature remains challenging for patients without computed tomography (CT) images that capture the abdomen or lower thorax.

**Objectives:** To (1) define sarcopenia thresholds at the C3 vertebral level using previously established thresholds derived from abdominal CT imaging and (2) determine if C3-defined sarcopenia is associated with survival in patients with HNC.

**Design, setting, and participants:** This retrospective cohort study was conducted in consecutive patients with a squamous cell carcinoma of the head and neck with cross-sectional abdominal or neck imaging within 60 days prior to treatment and treated between January 2005 and December 2017. Data analysis was completed from December 2018 to April 2021.

**Exposures:** Measurement of the cross-sectional muscle area at the third lumbar and cervical vertebral levels using CT imaging.

**Main outcomes and measures:** Primary study outcome was overall survival.

**Results:** In a cohort of 253 HNC patients with CT imaging that captures both L3 and C3 vertebral levels, skeletal muscle cross-sectional area at C3 was strongly correlated with the L3 level in both men (n = 188; r = 0.77; p < 0.001) and women (n = 65; r = 0.80; p < 0.001), and C3-defined sarcopenia thresholds of 14.0 cm^2^/m^2^ (men) and 11.1 cm^2^/m^2^ (women) were best predictive of previously established L3-defined sarcopenia thresholds. Applying these defined C3 sarcopenia thresholds in a cohort of 536 HNC patients with neck imaging alone revealed that C3-defined sarcopenia was independently associated with reduced overall survival in men (HR = 2.63; 95% CI, 1.79, 3.85) but not women (HR = 1.18, 95% CI, 0.76, 1.85) with HNC.

**Conclusions and relevance:** This study identifies sarcopenia thresholds at the C3 level that best predict L3-defined sarcopenia in both men and women. In HNC, C3-defined sarcopenia is associated with poor survival outcomes in men, but not women, suggesting sarcopenia may differentially affect men and women with HNC.

**Key Points:** *Question:* Is cervical vertebrae muscle wasting a reliable predictor of sarcopenia-related mortality in patients with head and neck cancer (HNC)?

*Findings:* (1) We established sex-specific sarcopenia thresholds (men, 14.0 cm^2^/m^2^; women, 11.1 cm^2^/m^2^) at the third cervical vertebrae (C3) level through statistical modelling that correlates with previously established lumbar thresholds. (2) Applying these defined C3 sarcopenia thresholds in a large cohort of HNC patients with neck imaging alone revealed that C3-defined sarcopenia was associated with reduced overall survival in men (HR = 2.63; 95% CI, 1.79, 3.85), but not women (HR = 1.18, 95% CI, 0.76, 1.85) with HNC.

*Meaning:* The C3 sarcopenia thresholds established herein may be a useful prognostic and risk-stratification tool; the influence of sarcopenia on patient outcomes should be assessed in a sex-specific manner.

## Introduction

Patients with cancer frequently experience weight loss, including progressive lean and fat mass catabolism consistent with the paraneoplastic wasting syndrome of cachexia^1,2^. Excessive skeletal muscle wasting, or sarcopenia, is significantly associated with morbidity and mortality for patients with solid tumors^3-5^. While sarcopenia is richly described as a negative prognostic marker for patients with primary tumors in the abdomen, only recently has its importance been identified in patients with head and neck cancer (HNC)^6-8^. Indeed, sarcopenia is a significant predictor of survival and post-operative complications for patients with HNC, and shows promise as a risk-stratification tool for patients with this disease^6-9^. Despite the clear utility in identifying sarcopenia in patients with HNC, clinical implementation of sarcopenia measures remains challenging due to the lack of established approaches that integrate with standard clinical workflow^10^.

Current skeletal muscle index (SMI) values for defining sarcopenia are specific to abdominal or lower thoracic musculature^5,11^. As abdominal and lower thoracic imaging is not always performed as part of the HNC workup, only a subset of patients that have imaging studies capturing these regions are included in the majority of previous reports, introducing potential selection bias. Therefore, defining and validating sarcopenia thresholds from routinely acquired head and neck images would greatly enhance clinical implementation while allowing better cross-study comparisons. Previous reports demonstrated a significant correlation between cervical and abdominal vertebrae cross sectional muscle area in patients with HNC, suggesting that routine head and neck imaging could be used to identify sarcopenia^12-15^. In the present study, we sought to expand upon these findings by defining sex-specific sarcopenia thresholds and evaluating their prognostic value and validating these associations in an independent patient cohort.

## Methods

### Population Cohort and End Points

We performed a retrospective review by screening medical records of patients who underwent primary surgical resection of head and neck squamous cell carcinoma between January 1, 2005 and December 21, 2017 at Oregon Health and Science University in Portland, Oregon. For comparative analysis between lumbar and cervical vertebrae imaging (training cohort), patients were required to have whole body PET-CT scans within 60 days prior to surgical resection. After establishment of C3 sarcopenia thresholds that best predicted previously defined L3 sarcopenia thresholds^5^, a second cohort of patients which had imaging of the head and neck alone (validation cohort) within 60 days prior to surgical resection were evaluated. Electronic health records were reviewed for data collection and included: patient demographics, body mass and height, comorbidities, tumor staging and subsite information, HPV/p16 status, smoking status, treatment information, evidence of recurrence, date and cause of death, and date of last follow up. After patient data were abstracted and coupled to their imaging information, all patient data were de-identified for subsequent analyses. This study was approved by the Institutional Review Board at Oregon Health and Science University. Requirement for informed consent was waived due to the retrospective nature of this study.

### Computed Tomography Body Composition Analysis

Body composition analysis of skeletal muscle was performed as previously described in patients with HNC^8,12^. Briefly, the cross-sectional area of skeletal muscle at the center of the third lumbar (L3) and third cervical (C3) vertebrae was determined by segmentation of axial CT images (Figure 1). Segmentation analysis was performed using Slice-o-Matic Software (version 5.0; Tomovision) to define muscle tissue cross-sectional area. Muscles delineated in the segmentation analysis included the rectus abdominus, abdominal wall, psoas, and the paraspinal muscle groups at the L3 level. Muscles measured at C3 included the paraspinal muscle group and the sternocleidomastoid. Muscle tissue was defined as - 29 to 150 Hounsfield units as described previously^16^. The resulting cross-sectional muscle area was then normalized to the square of the patient’s height in meters and used to calculate skeletal muscle index (SMI). At the L3 level, sarcopenia is defined as an SMI of less than 52.4 cm^2^/m^2^ for men and 38.5 cm^2^/m^2^ for women^5^. These thresholds are consistent with previous reports in head and neck cancer patients.

**Figure 1.**
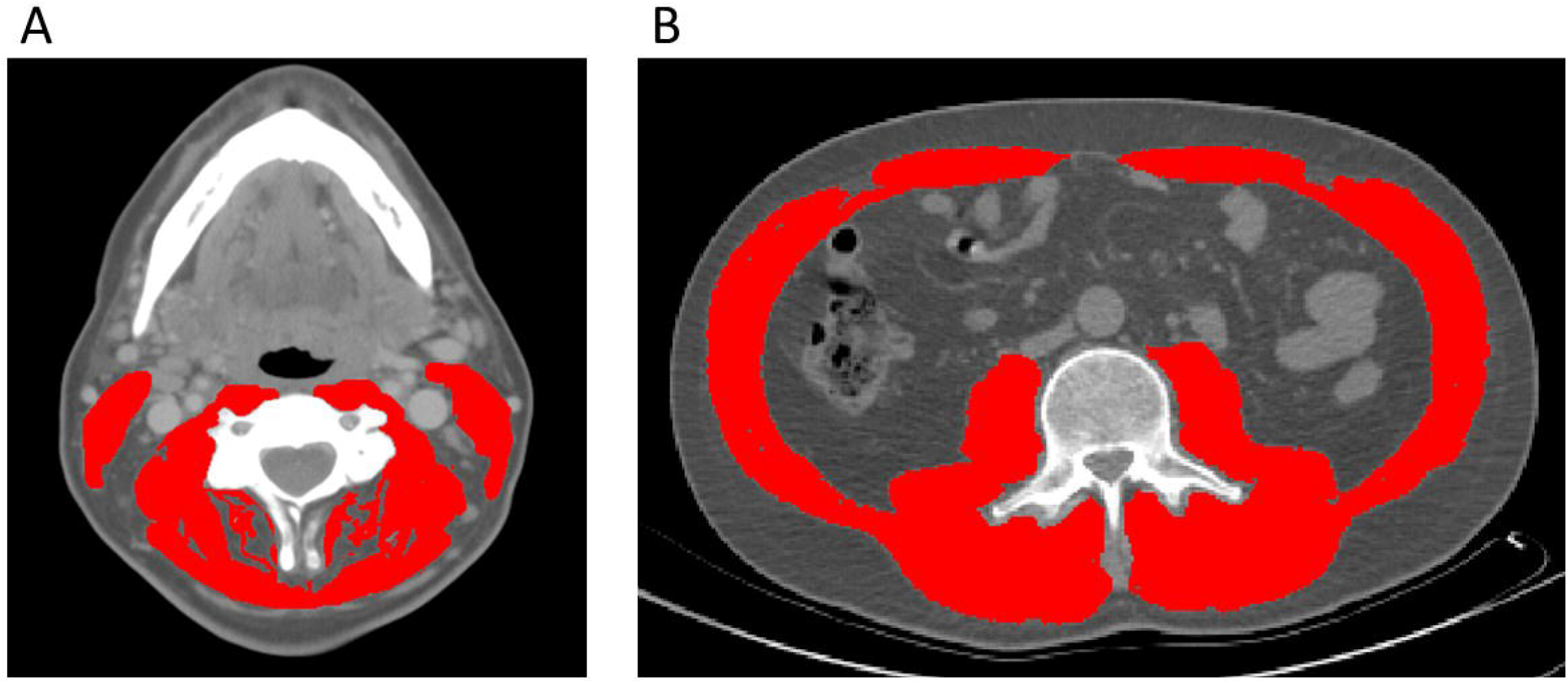
Representative axial CT images of the third lumbar (A) and third cervical (B) vertebral levels used to quantify skeletal muscle index (highlighted in red).

### Statistical Analysis

Data were analyzed between December 3, 2018, and April 28, 2021. To compare patient and clinical-disease characteristics at baseline, the Chi-square or Fisher’s Exact tests were used for categorical variables; the Wilcoxon rank sum test was used for continuous variables. Data was presented as median (IQR) or frequency (%). Training cohort (n = 253) comparisons were stratified by L3-defined sarcopenia; validation cohort (n = 536) comparisons were stratified by C3-defined sarcopenia.

Correlation was assessed using Pearson’s rho, which assumes normal distribution and a linear relationship between the measurements. Rho greater than 0.8 is considered ‘very strong’ correlation; rho between 0.6-0.79 suggests ‘strong’ correlation. To test for linearity and homoscedasticity we plotted residuals versus fitted values, showing the loess (smoothing) curve in red. Receiver Operating Characteristic (ROC) curves were generated to show the general predictive ability of C3 to predict L3-defined sarcopenia; DeLong’s test of correlated ROC curves was used to discriminate between C3 measurement types^17^. We used Youden’s Index to determine the optimal C3 cut-off value for predicting sarcopenia.

Overall survival is the time from initial diagnosis until death by any cause, with participants censored at their last assessment date. Assumptions of proportionality in the survival models were verified graphically and using residual-based models. Univariate and multivariable Cox proportional hazard ratio models were used to assess the risk of death based on demographic and baseline clinical-disease characteristics. We used purposeful selection combined with Bayesian information criterion (BIC) to build the multivariable models, entering all variables from the univariate models with p-value < 0.2. Kaplan-Meier curves with log-rank test were used to display overall survival stratified by C3-defined sarcopenia. All analyses were conducted using R, version 3.5.3.

## Results

### Determination of C3-defined sarcopenia thresholds

The training cohort included patients with both abdominal and neck imaging in order to perform intrapatient correlative analyses of L3 and C3 SMI values to identify appropriate C3 sarcopenia thresholds. Median age of this cohort is 61 (IQR 54, 68) years with 188/253 (74%) patients identifying as male. Patients in this population were classified as underweight (BMI <18.5; 15 [6.0%]), normal weight (BMI 18.5-24.9; 82 [32%]), overweight (BMI 25-29.9; 95 [38%]), or obese (BMI >30; 60 [24%]). Patients’ smoking status was binned in 3 groups, including 89 (35%) never smokers, 51 (20%) patients with <10 pack years, and 113 (45%) patients with ≥10 pack years. Postoperatively, 130 (51%) patients received temporary feeding tubes while 47 (19%) patients still had feeding tubes *in situ* at the time of last follow up. The majority of this cohort had oropharynx disease (147 patients [58%]), while primary tumors in the oral cavity (55 [22%]) and larynx (19 [8%]) were less frequently observed. Charlson Comorbidity Index (CCI) score was calculated as previously described^18^ and patients were subsequently stratified as either low risk (CCI <5; 213 [84%]) or high risk (CCI ≥5; 40 [16%]). One hundred three (41%) patients were treated by primary surgical resection alone, while 150 (59%) patients received adjuvant therapy (radiation and/or chemotherapy). Full characteristics and details of this cohort stratified by L3-defined sarcopenia are shown in eTable 1 in the Supplement.

Intrapatient L3 and C3 levels were strongly correlated in both men (n = 188, r = 0.77; p < 0.001) and women (n = 65, r = 0.80; p < 0.001; Figure 2; Supplemental Figure 1). As it is possible for the borders of the SCM to be obscured by lymph node metastases, we performed comparative analyses amongst L3 and C3 SMI values, both inclusive and exclusive of the SCM, to examine whether inclusion of the SCM improves or worsens predictive capacity of L3-based sarcopenia. Including the SCM in C3 SMI measurements improved predictive capacity of L3-defined sarcopenia in women (AUC = 89.4% vs. 86.3%; p = 0.03), but not men (AUC = 85.9% vs. 85.0%; p = 0.30; Supplemental Figure 2). Therefore, we included the SCM in all subsequent analyses. The C3 SMI thresholds with the best model performance based on Youden’s Index were 14.0 cm^2^/m^2^ for men and 11.1 cm^2^/m^2^ for women (Figure 2).

**Figure 2.**
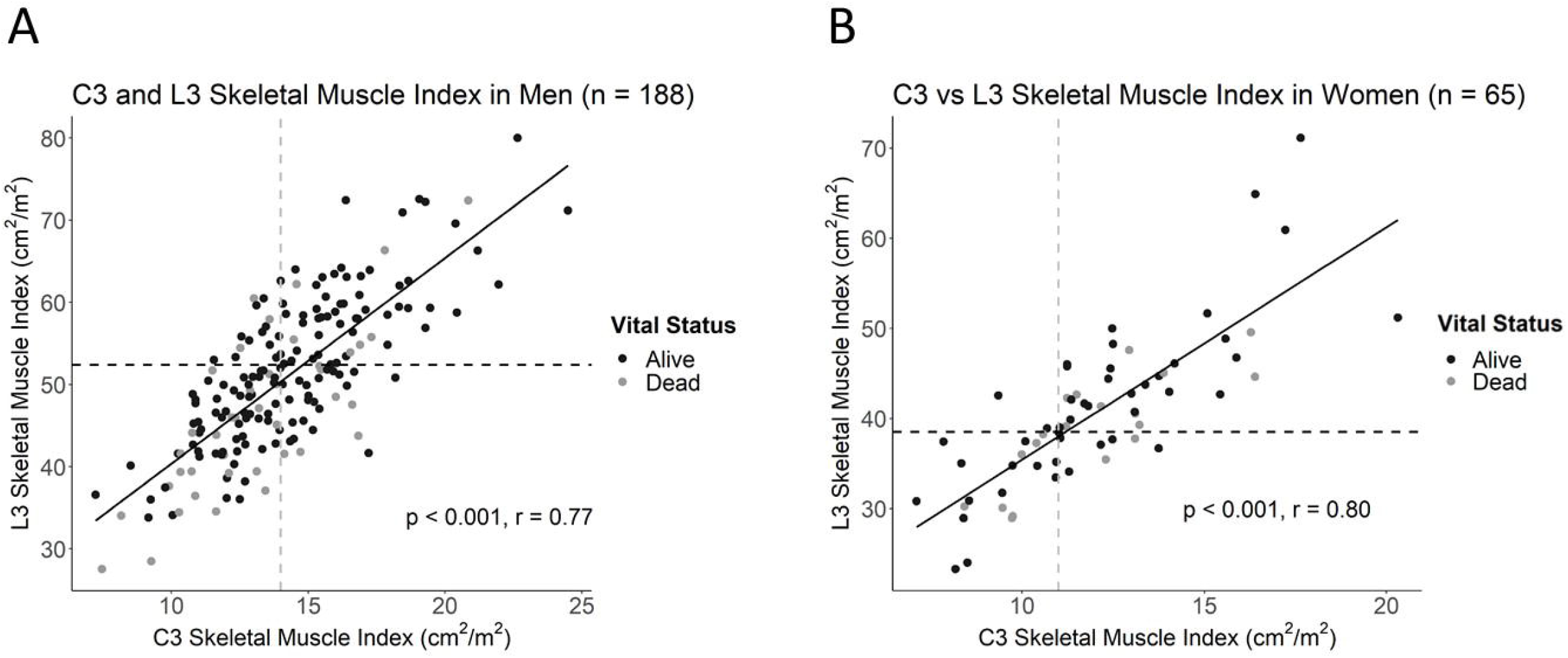
Correlation plots of L3 and C3 skeletal muscle indices in (A) men and (B) women with head and neck cancer. Dotted lines delineate the intersection of previously established L3 sarcopenia thresholds with the estimated C3 sarcopenia thresholds.

### Survival analyses in C3-defined sarcopenic patients

We next applied these C3 SMI thresholds to an independent cohort of HNC patients with imaging studies (PET-CT or CT) that captured the head and neck, but not abdomen or lower thorax (Table 1). This validation cohort included patients with imaging of the head and neck (but not abdomen). Median age of patients in this cohort was 64 (IQR 56, 72) with 333 patients identifying as male (62%) and 203 identifying as female (38%; Table 1). 41 (7.7%) patients were underweight, 211 (40%) were normal weight, 164 (31%) were overweight, and 118 (22%) were classified as obese (BMI >30). This cohort included 162 (30%) never smokers, 68 (13%) patients with <10 pack years, and 306 (57%) patients with ≥10 pack years. Postoperatively, 174 (32%) patients received temporary feeding tubes and 105 (20%) patients still had feeding tubes at last follow up. In contrast to the training cohort, the majority of this cohort had oral cavity disease (306 patients [57%]), while primary tumors in the oropharynx (118 [22%]) and larynx (79 [15%]) were less frequently observed. Using CCI, 409 [76%] patients were classified as low risk (CCI <5) and 127 were classified as [24%] high risk (CCI ≥5). In the validation cohort, 324 (60%) patients were treated by primary surgical resection alone, with 212 (40%) receiving adjuvant therapy (radiation and/or chemotherapy). Baseline patient characteristics stratified by C3-defined sarcopenia are shown in Table 1.

**Table 1:**
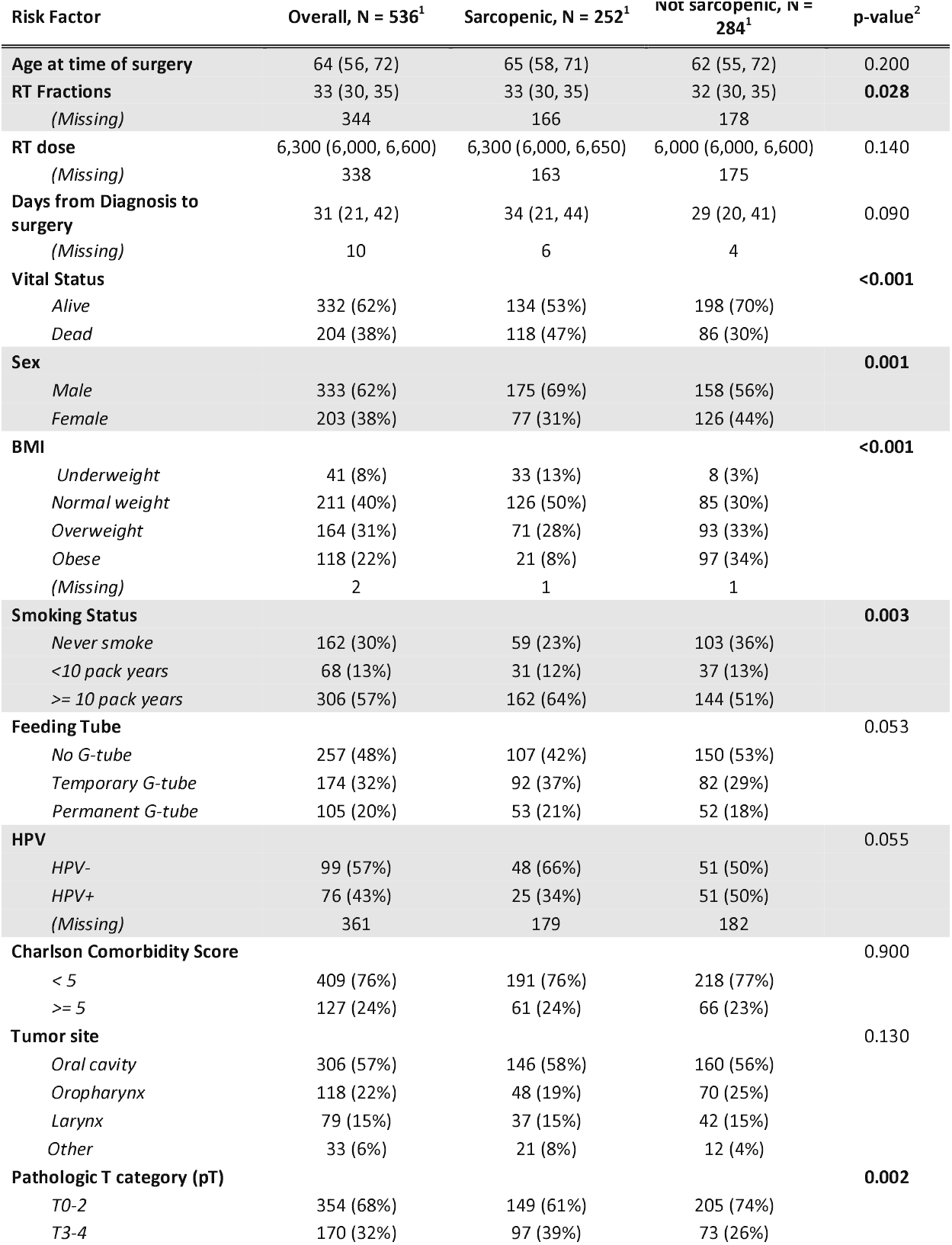
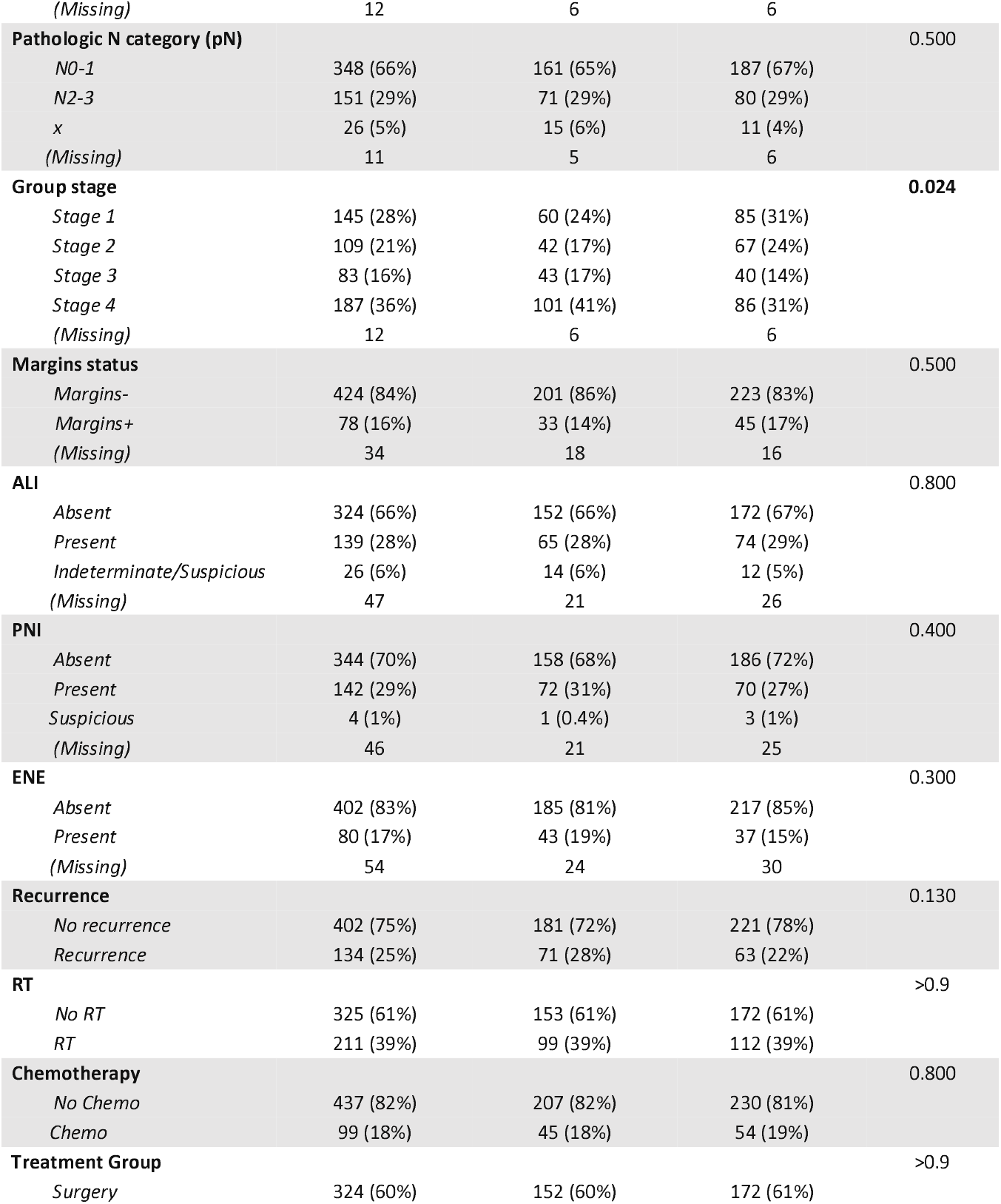
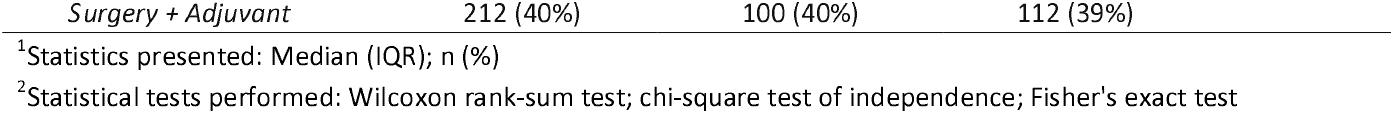
Clinical-demographic tables for validation set, stratified by C3-defined sarcopenia [Men: C3<14.0; Women: C3<11.1; n = 536)

In this cohort, 53% (175/333) of men were classified as sarcopenic compared to just 38% (77/203) of women (Table 1). Univariate Cox proportional hazards analysis revealed advanced age at time of surgery, C3-defined sarcopenia, underweight BMI status, >10 pack-year smoking status, permanent feeding tube placement, negative HPV status, elevated CCI, pT3-4, pN2-3, group stage 4, angiolymphatic invasion (ALI), perineural invasion (PNI), and extranodal extension (ENE) as significant predictors of survival in men. Univariate cox modelling for variables associated with survival in women included permanent feeding tube placement, elevated CCI, T category (pT3-4), group stage 3 and 4, ALI, and PNI (eTable3). Kaplan-Meier survival curves showed a significant difference in survival based on C3-defined sarcopenia for men but not women (Figure 3). On multivariable analyses, C3-defined sarcopenia (HR = 2.67; 95% CI, 1.72-4.15), ALI (HR = 2.00; 95% CI, 1.32-3.02), permanent feeding tube placement (HR = 2.33; 95% CI, 1.41-3.88), and age at time of surgery (HR = 1.04; 95% CI, 1.02-1.05) all remained significantly associated with overall survival for men (Table 2). In women, PNI (HR = 2.45; 95% CI, 1.48-4.06), ALI (HR = 2.13; 95% CI, 1.22-3.72), and pathologic T category (HR = 1.95; 95% CI, 1.20-3.16) were associated with reduced overall survival on multivariable analysis (Table 2).

**Figure 3.**
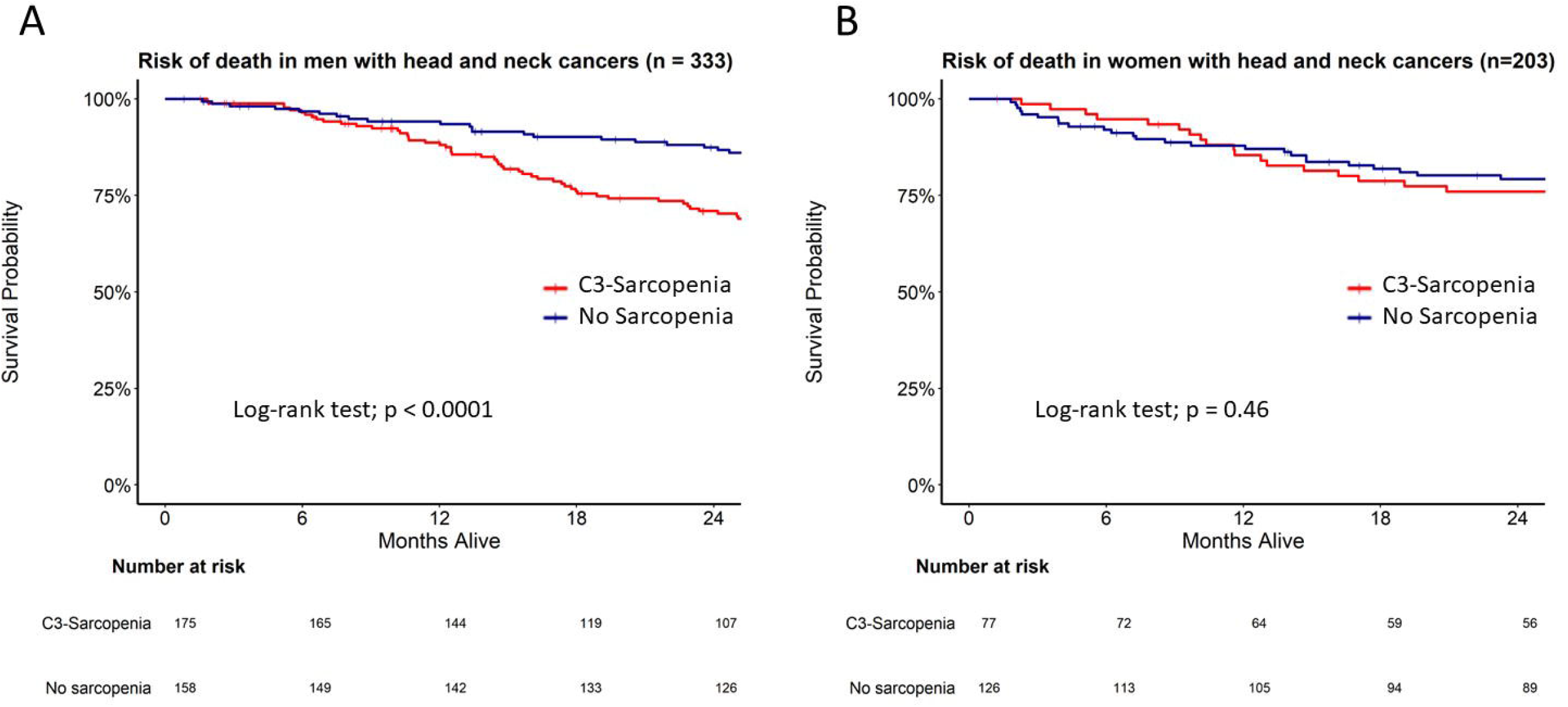
Survival analysis in C3-defined sarcopenic male and female HNC patients. Kaplan Meier curves for (A) men and (B) women stratified by C3 sarcopenia.

**Table 2:** Multivariable model for factors associated with risk of death with head and neck cancers in validation cohort, by sex (n = 536)

|  | Model selected for Men<br>(n = 333) |  |  | Model selected for Women<br>(n = 203) |  |  |
| --- | --- | --- | --- | --- | --- | --- |
| Variables: | HR<sup>1</sup> | 95% CI<sup>1</sup> | p-value | HR<sup>1</sup> | 95% CI<sup>1</sup> | p-value |
| **C3-Sarcopenia** |  |  | **<0.001** |  |  |  |
| Not sarcopenic | — | — |  |  |  |  |
| Sarcopenic | 2.67 | 1.72, 4.15 |  |  |  |  |
| **ALI** |  |  | **0.001** |  |  | **0.011** |
| Absent | — | — |  | — | — |  |
| Present | 2.00 | 1.32, 3.02 |  | 2.13 | 1.22, 3.72 |  |
| **Feeding Tube** |  |  | **0.002** |  |  |  |
| No G-tube | — | — |  |  |  |  |
| Temporary G-tube | 1.09 | 0.66, 1.81 |  |  |  |  |
| Permanent G-tube | 2.33 | 1.41, 3.88 |  |  |  |  |
| **Age at time of surgery** | 1.04 | 1.02, 1.05 | **<0.001** |  |  |  |
| **PNI** |  |  |  |  |  | **<0.001** |
| Absent |  |  |  | — | — |  |
| Present |  |  |  | 2.45 | 1.48, 4.06 |  |
| **Pathologic T category (pT)** |  |  |  |  |  | **0.009** |
| T0-2 |  |  |  | — | — |  |
| T3-4 |  |  |  | 1.95 | 1.20, 3.16 |  |

<sup>1</sup>HR = Hazard Ratio, CI = Confidence Interval

## Discussion

Skeletal muscle depletion is a well-established prognostic marker in multiple clinical and disease settings, including cancer^6,7^, trauma^19^, and scaling drug dosing to minimize toxicities^20^. Previous reports demonstrate the prognostic utility of sarcopenia is independent of patient body mass^5,7,21^, and that height and weight formulae are not sufficient to capture sarcopenia^22^. However, identification of this high-risk disease feature is challenging in HNC because imaging studies that capture the abdomen or lower thorax are infrequent in this population. Although previous studies demonstrate promise in utilizing neck imaging as a marker of sarcopenia^12,15,23^, we sought to expand upon these foundational studies by putting forth normalized sarcopenia thresholds and testing their prognostic utility in an independent cohort of patients with neck imaging alone. To that end, we identified 253 HNC patients with imaging studies that captured both the abdomen and neck and performed correlative analyses to identify sex-specific C3 sarcopenia thresholds that best predicted previously established and validated L3 SMI sarcopenia thresholds^5^. We then applied these C3-defined thresholds to an independent cohort of 536 HNC patients with imaging studies that only captured neck musculature to determine if our newly defined C3 sarcopenia thresholds were associated with survival outcomes. To our knowledge, our study is the first to establish normalized sex-specific C3 sarcopenia thresholds and validate their prognostic value in an independent cohort of HNC patients with imaging studies specific to the neck. We found that these C3 SMI thresholds were prognostic in men, but not women, with HNC.

Several foundational studies demonstrated a strong correlation of L3 and C3 skeletal muscle area in patients with HNC consistent with our findings herein^12,15,24,25^. Abdominal skeletal muscle cross-sectional area is linearly related to total body muscle mass, so we normalized C3 muscle cross sectional area for stature to determine sex-specific thresholds for sarcopenia as previously described^5^. Our study reveals C3 sarcopenia thresholds of 14.0 cm^2^/m^2^ and 11.1 cm^2^/m^2^ in men and women, respectively, best predict previously established and widely published L3 sarcopenia thresholds^5^. Although recent work utilized cross-sectional muscle area at the C3 level to predict L3 cross-sectional area, the authors classified sarcopenia as the lowest quartile of predicted L3 muscle area, while the work herein sought to base C3 sarcopenia thresholds on L3 sarcopenia thresholds that are validated across multiple cancers and conditions^15^. Accordingly, it is possible that these C3 sarcopenia thresholds extend to other diseases and clinical settings in which the abdomen or lower thorax are not routinely captured by CT imaging, such as neurologic cancers. However, identifying the most suitable sarcopenia thresholds appears to be multifactorial and dependent on sex and disease-specific conditions^26^, further highlighting the need for validating sarcopenia thresholds in a context-specific manner.

As described above, when we apply these thresholds to a large cohort of HNC patients stratified by sex, C3-defined sarcopenia at the time of diagnosis was associated with reduced survival in men, but not women. To our knowledge, this is the largest dataset of women with HNC stratified by sarcopenia, and the first to individually assess sex-specific survival outcomes during this disease. Like many research areas, sarcopenia has largely been studied in males in both murine and clinical settings, while few studies evaluating sex as a biological variable exist^27,28^. Amongst experimental rodent models of colon- and HNC-associated cachexia, male mice lost a larger proportion of lean mass compared to female mice with similar disease burden^29,30^. Several studies also report a significantly lower prevalence of sarcopenia in women with various cancers, including non-small cell lung and gastrointestinal cancers^31 32^. In the present study, we observe 55% of men are sarcopenic at the time of diagnosis compared to just 39% of women (p < 0.001). In addition to reports that describe a decrease in the prevalence of sarcopenia in women at the time of cancer diagnosis, Kilgour and colleagues showed a strong association between muscle mass and cancer-associated fatigue in men, but not in women^33^. Burkart and colleagues recently showed an association between sarcopenia and overall survival in men with aggressive B-cell Non-Hodgkin Lymphoma, while women demonstrated no such association^34^. Conversely, sarcopenia was associated with poorer survival outcomes in women with non-metastatic breast cancer^21^. These studies and more suggest that sarcopenia may not only disproportionately appear between men and women during disease, but also differentially influence quality of life and mortality in a sex- and disease-specific manner^26^.

It is plausible that the sarcopenia observed in this population is associated with cachexia, a disease-associated metabolic syndrome that significantly reduces patient’s quality of life and ultimate survival^1^. In the present study, it is possible that sarcopenia portends the development of cachexia more frequently in men than women, or that muscle loss is not associated with impaired resilience in women with HNC—future prospective investigations are needed to test these hypotheses. While this study demonstrates pre-therapy C3-defined sarcopenia is a useful prognostic marker in men, but not women, diagnosed with HNC, a recent report demonstrated that conversion to sarcopenia (detected by L3 muscle area) was associated with a reduction in overall survival in patients with HNC treated with definitive RT ^6^. Therefore, the sex-specific influence of post-therapy conversion to sarcopenia using the C3-sarcopenia thresholds established herein remains an area of active investigation. Collectively, our findings highlight the need for future studies to be deliberate in examining sex-specific effects and prevalence of sarcopenia.

### Limitations

This study has several limitations that should be taken into consideration when interpreting these data. As with any retrospective review, this study is subjected to missing data and heterogeneous patient follow-up. This study was performed at a single tertiary care institution, resulting in a patient demographic that may not be reflective of other areas of the country. Finally, given the timeframe of this study that traverses the recognition of HPV/p16 as a prognostic factor, these patients are staged by the American Joint Committee on Cancer (AJCC) seventh edition, as opposed AJCC eight edition staging criteria.

C3 thresholds were not developed against the ‘gold standard’ for sarcopenia, either volumetric full body cross sectional imaging or dual x-ray absorptiometry. A limitation of this study is that a surrogate marker of sarcopenia, normalized L3 skeletal muscle cross sectional area, is used to estimate the C3 threshold value and may imprecisely estimate sarcopenia. However, this measure is associated with survival endpoints in multiple disease states, supporting its use in the present study. We directly tested C3 measurements with risk of death using a partitioning approach (partDSA survival package in R) which generated a similar threshold value for men but provided no distinct C3 cut-off value for women.

## Conclusions

Taken together, this study demonstrates that C3-sarcopenia thresholds are strongly associated with previously defined L3 sarcopenia thresholds, and C3-defined sarcopenia is independently associated with reduced survival in men, but not women with HNC. We propose that the C3-defined sarcopenia thresholds herein represent a useful prognostic tool for men with HNC. Future research concerning these observations is warranted, including determining whether these thresholds and sex-specific survival associations extend to other pathologies.

## Data Availability

Data are available upon reasonable request.

## Acknowledgements

This work was supported National Cancer Institute grants CA245188 (Grossberg) and CA254033 (Olson). Dr. Fuller received/receives funding and salary support during the period of study execution unrelated to this work from: the National Institutes of Health (NIH) National Cancer Institute (NCI) Early Stage Development of Technologies in Biomedical Computing, Informatics, and Big Data Science Program (R01CA214825); Joint NSF/NIH Initiative on Quantitative Approaches to Biomedical Big Data program (R01CA225190); NIH National Institute of Biomedical Imaging and Bioengineering (NIBIB) Research Education Programs for Residents and Clinical Fellows Grant (R25EB025787); NIH National Institute of Dental and Craniofacial Research (NIDCR) Academic Industrial Partnership Grant (R01DE028290); NIDCR Establishing Outcome Measures for Clinical Studies of Oral and Craniofacial Diseases and Conditions award (R01DE025248); NCI Parent Research Project Grant (R01CA258827); NCI Early Phase Clinical Trials in Imaging and Image-Guided Interventions Program (1R01CA218148); an NIH/NCI Cancer Center Support Grant (CCSG) Pilot Research Program Award from the UT MD Anderson CCSG Radiation Oncology and Cancer Imaging Program (P30CA016672); a sub-award from the Small Business Innovation Research Grant Program (R43CA254559); a sub-award from the The Human BioMolecular Atlas Program (HuBMAP) Integration, Visualization & Engagement (HIVE) Initiative (OT2OD026675); the Patient-Centered Outcomes Research Institute (PCS-1609-36195); a National Science Foundation (NSF) Division of Civil, Mechanical, and Manufacturing Innovation (CMMI) grant (NSF 1933369); and the NSF/NCI Smart connected Health Program (R01CA257814). Dr. Fuller receives infrastructure support from MD Anderson Cancer Center under the The Image Guided Cancer Therapy (IGCT) Research Program, and grant and infrastructure support via the Charles and Daneen Stiefel Center for Head and Neck Cancer Oropharyngeal Cancer Research Program. Dr. Fuller has received direct industry grant/in-kind support, honoraria, and travel funding from Elekta AB unrelated to this project.

