## Supplemental materials for "Establishment and validation of pre-therapy cervical vertebrae muscle quantification as a prognostic marker of sarcopenia in head and neck patients receiving definitive cancer surgery"

Supplementary Online Content

eTable 1. Baseline Patient Characteristics of OHSU patients with imaging capturing the abdomen and neck, training cohort (men, n = 188, women, n = 65).

eTable 2. Body Composition and C3-defined sarcopenia status of OHSU patients with imaging capturing only the neck stratified by sex, validation cohort (men, n = 333; women, n=203)

eTable 3. Univariate Cox Models for variables associated with risk of death with head and neck cancers, by sex, validation cohort (men, n = 333; women, n=203). Reports Wald p-value.

eTable 1.

| **Variables** | **Overall, N = 253^1^** | **Sarcopenia, N = 140^1^** | **Not sarcopenic, N = 113^1^** | **p-value^2^** |
| --- | --- | --- | --- | --- |
| **Age** | 61 (54, 68) | 63 (58, 71) | 57 (52, 63) | **<0.001** |
| **RT Fractions** | 32 (30, 35) | 32 (30, 35) | 33 (30, 35) | 0.900 |
| *(Missing)* | 147 | 83 | 64 |  |
| **RT dose** | 6,300 (6,000, 6,400) | 6,300 (6,000, 6,400) | 6,060 (6,000, 6,532) | 0.700 |
| *(Missing)* | 123 | 70 | 53 |  |
| **C3-Sarcopenia** |  |  |  | **<0.001** |
| *Sarcopenia* | 124 (49%) | 105 (75%) | 19 (17%) |  |
| *No sarcopenia* | 129 (51%) | 35 (25%) | 94 (83%) |  |
| **Vital Status** |  |  |  | **0.046** |
| *Alive* | 192 (76%) | 99 (71%) | 93 (82%) |  |
| *Dead* | 61 (24%) | 41 (29%) | 20 (18%) |  |
| **Sex** |  |  |  | **0.031** |
| *Male* | 188 (74%) | 112 (80%) | 76 (67%) |  |
| *Female* | 65 (26%) | 28 (20%) | 37 (33%) |  |
| **BMI** |  |  |  | **<0.001** |
| *Underweight* | 15 (6.0%) | 14 (10%) | 1 (0.9%) |  |
| *Normal weight* | 82 (32%) | 62 (45%) | 20 (18%) |  |
| *Overweight* | 95 (38%) | 49 (35%) | 46 (41%) |  |
| *Obese* | 60 (24%) | 14 (10%) | 46 (41%) |  |
| *(Missing)* | 1 | 1 | 0 |  |
| **Smoking Status** |  |  |  | **0.003** |
| *Never smoke* | 89 (35%) | 40 (29%) | 49 (43%) |  |
| *<10 pack years* | 51 (20%) | 24 (17%) | 27 (24%) |  |
| *>= 10 pack years* | 113 (45%) | 76 (54%) | 37 (33%) |  |
| **Feeding Tube** |  |  |  | 0.130 |
| *No G-tube* | 76 (30%) | 38 (27%) | 38 (34%) |  |
| *Temporary G-tube* | 130 (51%) | 70 (50%) | 60 (53%) |  |
| *Permanent G-tube* | 47 (19%) | 32 (23%) | 15 (13%) |  |
| **HPV** |  |  |  | 0.120 |
| *HPV-* | 33 (27%) | 22 (33%) | 11 (19%) |  |
| *HPV+* | 90 (73%) | 44 (67%) | 46 (81%) |  |
| *(Missing)* | 130 | 74 | 56 |  |
| **Tumor site** |  |  |  | 0.130 |
| *Oral cavity* | 55 (22%) | 37 (27%) | 18 (16%) |  |
| *Oropharynx* | 147 (58%) | 76 (55%) | 71 (63%) |  |
| *Larynx* | 19 (8%) | 12 (9%) | 7 (6%) |  |
| *Other* | 31 (12%) | 14 (10%) | 17 (15%) |  |
| *(Missing)* | 1 | 1 | 0 |  |
| **Charlson Comorbidity Score** |  |  |  | **0.027** |
| *< 5* | 213 (84%) | 111 (79%) | 102 (90%) |  |
| *>= 5* | 40 (16%) | 29 (21%) | 11 (9.7%) |  |
| **Pathologic T category (pT)** |  |  |  | 0.300 |
| *T0-2* | 175 (70%) | 92 (67%) | 83 (73%) |  |
| *T3-4* | 76 (30%) | 46 (33%) | 30 (27%) |  |
| *(Missing)* | 2 | 2 | 0 |  |
| **Pathologic N category (pN)** |  |  |  | >0.9 |
| *N0-1* | 140 (56%) | 77 (56%) | 63 (56%) |  |
| *N2-3* | 111 (44%) | 61 (44%) | 50 (44%) |  |
| *(Missing)* | 2 | 2 | 0 |  |
| **Group stage** |  |  |  | 0.400 |
| *Stage 1* | 59 (24%) | 28 (20%) | 31 (27%) |  |
| *Stage 2* | 78 (31%) | 43 (31%) | 35 (31%) |  |
| *Stage 3* | 26 (10%) | 13 (9.4%) | 13 (12%) |  |
| *Stage 4* | 88 (35%) | 54 (39%) | 34 (30%) |  |
| *(Missing)* | 2 | 2 | 0 |  |
| **Margins status** |  |  |  | >0.9 |
| *Negative* | 194 (80%) | 108 (79%) | 86 (80%) |  |
| *Positive* | 50 (20%) | 28 (21%) | 22 (20%) |  |
| *(Missing)* | 9 | 4 | 5 |  |
| **ALI** |  |  |  | 0.400 |
| *Absent* | 119 (49%) | 62 (46%) | 57 (53%) |  |
| *Present* | 123 (51%) | 72 (54%) | 51 (47%) |  |
| *(Missing)* | 11 | 6 | 5 |  |
| **PNI** |  |  |  | 0.400 |
| *Absent* | 161 (66%) | 86 (64%) | 75 (69%) |  |
| *Present* | 82 (34%) | 49 (36%) | 33 (31%) |  |
| *(Missing)* | 10 | 5 | 5 |  |
| **ENE** |  |  |  | >0.9 |
| *Absent* | 167 (69%) | 92 (69%) | 75 (69%) |  |
| *Present* | 75 (31%) | 41 (31%) | 34 (31%) |  |
| *(Missing)* | 11 | 7 | 4 |  |
| **RT** |  |  |  | 0.400 |
| *No RT* | 105 (42%) | 62 (44%) | 43 (38%) |  |
| *RT* | 148 (58%) | 78 (56%) | 70 (62%) |  |
| **Treatment group** |  |  |  | 0.400 |
| *Surgery* | 103 (41%) | 61 (44%) | 42 (37%) |  |
| *Surgery + adjuvant* | 150 (59%) | 79 (56%) | 71 (63%) |  |
| **Recurrence** |  |  |  | 0.300 |
| *No recurrence* | 195 (77%) | 104 (74%) | 91 (81%) |  |
| *Recurrence* | 58 (23%) | 36 (26%) | 22 (19%) |  |
| ^1^Statistics presented: Median (IQR); n (%) | | | | |
| ^2^Statistical tests performed: Wilcoxon rank-sum test; chi-square test of independence | | | | |

eTable 2.

| **Risk Factor** | **Overall, N = 536^1^** | **Male, N = 333^1^** | **Female, N = 203^1^** | **p-value^2^** |
| --- | --- | --- | --- | --- |
| **BMI** |  |  |  | 0.140 |
| *Underweight* | 41 (7.7%) | 20 (6.0%) | 21 (10%) |  |
| *Normal weight* | 211 (40%) | 129 (39%) | 82 (40%) |  |
| *Overweight* | 164 (31%) | 111 (34%) | 53 (26%) |  |
| *Obese* | 118 (22%) | 71 (21%) | 47 (23%) |  |
| *(Missing)* | 2 | 2 | 0 |  |
| **C3-Sarcopenia** |  |  |  | **0.001** |
| *Sarcopenic* | 252 (47%) | 175 (53%) | 77 (38%) |  |
| *Not sarcopenic* | 284 (53%) | 158 (47%) | 126 (62%) |  |
| **C3 SMI+SCM (cm^2^/m^2^)** | 13.2 (11.5, 15.1) | 13.8 (12.3, 15.8) | 12.0 (10.3, 13.8) | **<0.001** |

eTable 3.

|  | **Univariate Models for Men (n = 333)** | | | | | **Univariate Models for Women (n = 203)** | | | | |
| --- | --- | --- | --- | --- | --- | --- | --- | --- | --- | --- |
| **Risk Factor** | **N** | **Event N** | **HR^1^** | **95% CI^1^** | **p-value** | **N** | **Event N** | **HR^1^** | **95% CI^1^** | **p-value** |
| **Age at time of surgery** | 333 | 123 | 1.04 | 1.02, 1.05 | **<0.001** | 203 | 80 | 1.00 | 0.99, 1.02 | 0.651 |
| **RT Fractions** | 143 | 50 | 1.00 | 1.00, 1.01 | 0.284 | 49 | 19 | 1.00 | 0.99, 1.01 | 0.826 |
| **RT dose** | 146 | 51 | 1.00 | 1.00, 1.00 | 0.492 | 52 | 20 | 1.00 | 1.00, 1.00 | 0.358 |
| **Days from Diagnosis to surgery** | 325 | 118 | 1.00 | 0.99, 1.01 | 0.654 | 201 | 79 | 1.00 | 0.99, 1.01 | 0.457 |
| **C3-Sarcopenia** |  | 123 |  |  | **<0.001** |  | 80 |  |  | 0.465 |
| Not sarcopenic | 158 |  | — | — |  | 126 |  | — | — |  |
| Sarcopenic | 175 |  | 2.63 | 1.79, 3.85 |  | 77 |  | 1.18 | 0.76, 1.85 |  |
| **BMI** |  | 123 |  |  | **<0.001** |  | 80 |  |  | 0.16 |
| Normal weight | 129 |  | — | — |  | 82 |  | — | — |  |
| Underweight | 20 |  | 1.87 | 1.00, 3.49 |  | 21 |  | 1.68 | 0.87, 3.25 |  |
| Overweight | 111 |  | 0.57 | 0.37, 0.86 |  | 53 |  | 0.72 | 0.40, 1.29 |  |
| Obese | 71 |  | 0.47 | 0.28, 0.80 |  | 47 |  | 0.81 | 0.45, 1.46 |  |
| **Smoking Status** |  | 123 |  |  | **<0.001** |  | 80 |  |  | 0.488 |
| Never smoke | 88 |  | — | — |  | 74 |  | — | — |  |
| <10 pack years | 36 |  | 0.91 | 0.41, 2.03 |  | 32 |  | 0.78 | 0.36, 1.65 |  |
| >= 10 pack years | 209 |  | 2.10 | 1.33, 3.31 |  | 97 |  | 1.17 | 0.72, 1.88 |  |
| **Feeding Tube** |  | 123 |  |  | **<0.001** |  | 80 |  |  | **0.003** |
| No G-tube | 142 |  | — | — |  | 115 |  | — | — |  |
| Temporary G-tube | 116 |  | 1.35 | 0.87, 2.07 |  | 58 |  | 1.06 | 0.62, 1.79 |  |
| Permanent G-tube | 75 |  | 2.71 | 1.75, 4.20 |  | 30 |  | 2.7 | 1.57, 4.66 |  |
| **HPV** |  | 123 |  |  | **0.003** |  | 80 |  |  | **0.087** |
| HPV+ | 52 |  | -- | -- |  | 24 |  | -- | -- |  |
| HPV- | 67 |  | 2.44 | 1.09, 5.49 |  | 32 |  | 1.19 | 0.44,3.22 |  |
| Unknown | 214 |  | 2.93 | 1,42, 6.05 |  | 147 |  | 1.96 | 0.90, 4.29 |  |
| **Tumor site** |  | 123 |  |  | 0.205 |  | 80 |  |  | 0.401 |
| Oral cavity | 163 |  | — | — |  | 143 |  | — | — |  |
| Oropharynx | 89 |  | 0.68 | 0.43, 1.08 |  | 29 |  | 0.83 | 0.44, 1.58 |  |
| Larynx | 54 |  | 1.07 | 0.66, 1.73 |  | 25 |  | 0.54 | 0.25, 1.19 |  |
| Other | 27 |  | 1.29 | 0.69, 2.39 |  | 6 |  | 1.14 | 0.36, 3.66 |  |
| **Charlson Comorbidity Score** |  | 123 |  |  | **0.005** |  | 80 |  |  | **0.03** |
| < 5 | 259 |  | — | — |  | 150 |  | — | — |  |
| >= 5 | 74 |  | 1.78 | 1.21, 2.63 |  | 53 |  | 1.69 | 1.07, 2.68 |  |
| **Pathologic T category (pT)** |  | 121 |  |  | **<0.001** |  | 77 |  |  | **0.002** |
| T0-2 | 208 |  | — | — |  | 146 |  | — | — |  |
| T3-4 | 118 |  | 1.92 | 1.34, 2.75 |  | 52 |  | 2.13 | 1.34, 3.39 |  |
| **Pathologic N category (pN)** |  | 121 |  |  | **0.011** |  | 77 |  |  | 0.336 |
| N0-1 | 203 |  | — | — |  | 145 |  | — | — |  |
| N2-3 | 111 |  | 1.77 | 1.22, 2.56 |  | 40 |  | 1.52 | 0.89, 2.59 |  |
| x | 13 |  | 1.49 | 0.68, 3.26 |  | 13 |  | 1.14 | 0.45, 2.85 |  |
| **Group stage** |  | 121 |  |  | **<0.001** |  | 77 |  |  | **0.002** |
| Stage 1 | 79 |  | — | — |  | 66 |  | — | — |  |
| Stage 2 | 67 |  | 0.84 | 0.43, 1.64 |  | 42 |  | 1.32 | 0.65, 2.69 |  |
| Stage 3 | 49 |  | 1.39 | 0.74, 2.62 |  | 34 |  | 2.87 | 1.50, 5.48 |  |
| Stage 4 | 113 |  | 2.39 | 1.48, 3.87 |  | 56 |  | 2.60 | 1.43, 4.73 |  |
| **Margins status** |  | 110 |  |  | 0.182 |  | 79 |  |  | 0.914 |
| Margins- | 246 |  | — | — |  | 178 |  | — | — |  |
| Margins+ | 59 |  | 1.36 | 0.88, 2.11 |  | 19 |  | 1.04 | 0.50, 2.17 |  |
| **ALI** |  | 109 |  |  | **0.002** |  | 77 |  |  | **0.002** |
| Absent | 177 |  | — | — |  | 147 |  | — | — |  |
| Present | 106 |  | 1.85 | 1.25, 2.73 |  | 33 |  | 2.64 | 1.57, 4.45 |  |
| Indeterminate/Suspicious | 13 |  | 2.61 | 1.18, 5.77 |  | 13 |  | 0.72 | 0.22, 2.29 |  |
| **PNI** |  | 110 |  |  | **0.015** |  | 77 |  |  | **<0.001** |
| Absent | 208 |  | — | — |  | 136 |  | — | — |  |
| Present | 88 |  | 1.64 | 1.11, 2.42 |  | 54 |  | 2.66 | 1.68, 4.21 |  |
| **ENE** |  | 108 |  |  | **0.041** |  | 75 |  |  | 0.833 |
| Absent | 236 |  | — | — |  | 166 |  | — | — |  |
| Present | 57 |  | 1.62 | 1.04, 2.52 |  | 23 |  | 1.08 | 0.54, 2.17 |  |
| **Recurrence** |  | 123 |  |  | **<0.001** |  | 80 |  |  | **<0.001** |
| No recurrence | 253 |  | — | — |  | 149 |  | — | — |  |
| Recurrence | 80 |  | 2.37 | 1.65, 3.39 |  | 54 |  | 3.91 | 2.52, 6.09 |  |
| **RT** |  | 123 |  |  | 0.748 |  | 80 |  |  | 0.706 |
| No RT | 179 |  | — | — |  | 146 |  | — | — |  |
| RT | 154 |  | 0.94 | 0.66, 1.35 |  | 57 |  | 1.10 | 0.68, 1.77 |  |
| **Chemotherapy** |  | 123 |  |  | 0.827 |  | 80 |  |  | 0.819 |
| No Chemo | 261 |  | — | — |  | 176 |  | — | — |  |
| Chemo | 72 |  | 1.05 | 0.68, 1.61 |  | 27 |  | 1.08 | 0.57, 2.04 |  |
| **Treatment Group** |  | 123 |  |  | 0.748 |  | 80 |  |  | 0.554 |
| Surgery | 179 |  | — | — |  | 145 |  | — | — |  |
| Surgery + Adjuvant | 154 |  | 0.94 | 0.66, 1.35 |  | 58 |  | 1.16 | 0.72, 1.85 |  |
| ^1^HR = Hazard Ratio, CI = Confidence Interval | | | | | |  |  |  |  |  |

Supplementary Figure 1. Residual vs fitted value plots for men (n = 188) and women (n = 65). Loess curve is shown in red.

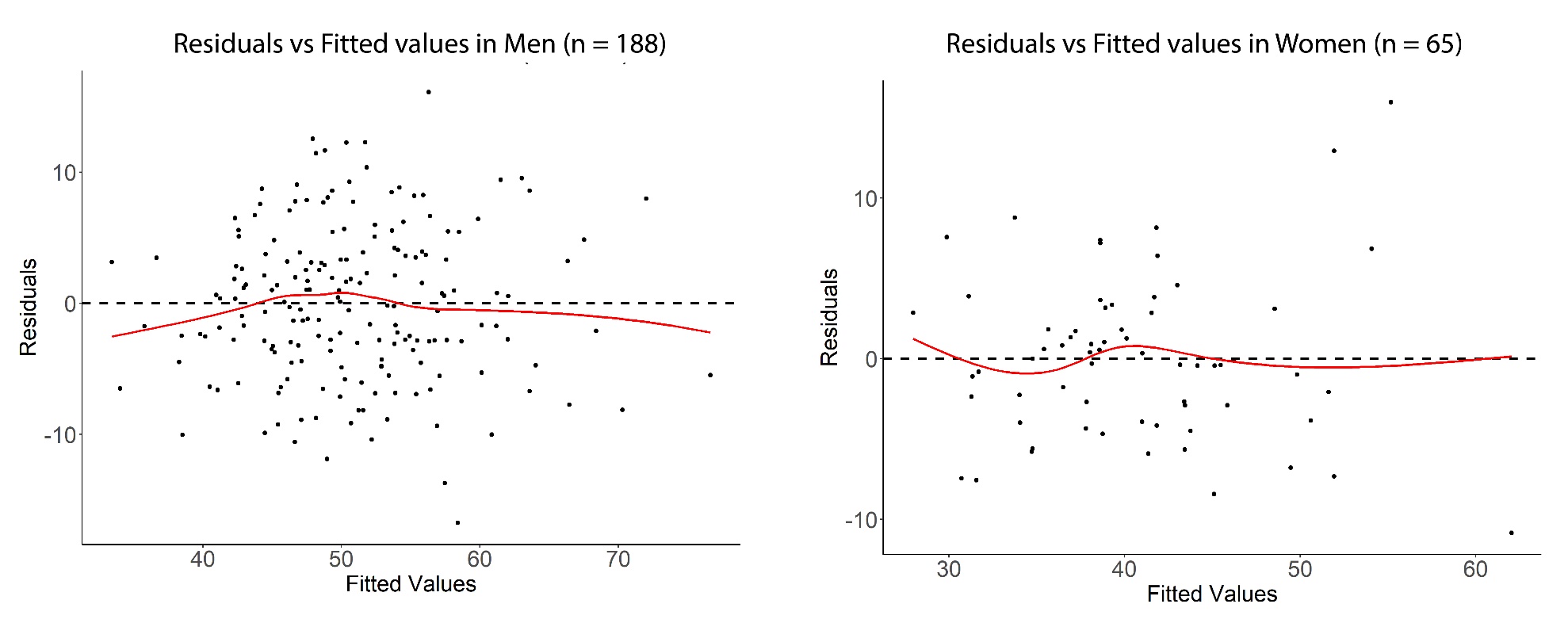

Supplementary Figure 2. ROC curves for L3 and C3 (with and without SCM) in men (n = 188) and women (n = 65).

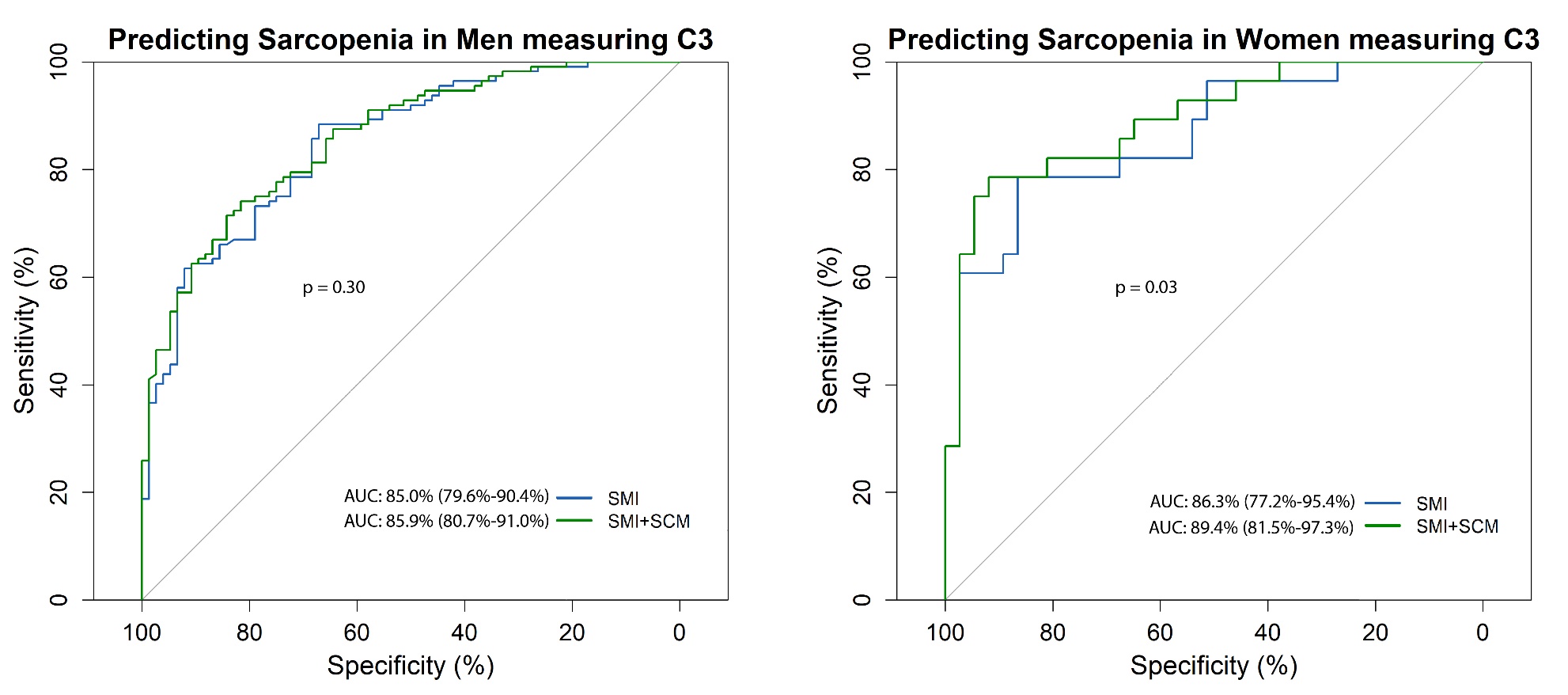
